## appendix 1 for "Further development and feasibility randomised controlled trial of a digital programme for adolescent depression, MoodHwb: study protocol"

**Proposed progression criteria – 'Digital support for young people with their mood and wellbeing' study**

| CRITERIA | INDICATOR<br>GREEN = very strong indication to proceed<br>AMBER = medium indication to proceed. Discuss with Trial Steering Committee (TSC) and proceed with identified plan.<br>RED = indication of doubt as to whether to proceed. Discuss with TSC, and only proceed if other indicators are amber/green and there is a clear mitigation strategy. | METHOD OF ASSESSMENT |
|---|---|---|
| **1. Feasibility and acceptability outcomes related to the trial methods** | | |
| 1.1. Recruitment rate | Actual recruitment rate vs. Target recruitment rate:<br>Green: ≥85% of participants<br>Amber: 60-84%<br>Red: <60% | Number of eligible young people who consent to participate in the study within the first 6 months of recruitment<br>Vs. Target number (120) within this period<br><br>[Also to review % of young people who showed an interest in the study who go on to participate] |
| 1.2. Retention rate | Retention rates:<br>Green: ≥75% of participants<br>Amber: 50-74%<br>Red: <50% | Number of young people who remained in the study at 2 months<br>Vs. Total number who consented to participate at baseline |
| 1.3. Completeness of outcome measures | Completion of core measures at baseline and follow-up:<br>Green: ≥90% data completion | % of participants who completed the core questionnaires (including measures on |

| | | |
|---|---|---|
| | Amber: 70-89%<br>Red: <70% | depressive and anxiety symptoms, wellbeing, knowledge and help-seeking)<br>% of missing data from completed core questionnaires<br><br>Both at i) baseline and ii) 2-month follow-up (i.e. based on completion by those who remained in the study)<br><br>[Also to review views/acceptability of trial methods from questionnaire, interview and focus group data] |
| **2. Feasibility and acceptability outcomes related to the digital programme** | | |
| 2.1. Level of usage of programme | % of participants in the intervention arm who accessed and used the programme:<br>Green: ≥80%<br>Amber: 60-79%<br>Red: <60%<br><br>Also to discuss number of times the programme has been accessed, duration of use and the sections/components accessed. | Web/app usage data from Google Analytics, based on log-ins into and use of MoodHwb by young people (and parents/carers) – up to 2 and 6 months from baseline<br><br>Questionnaire items on usage completed by young people and parents/carers at 2-month follow-up<br><br>Semi-structured interviews with young people and parents/carers at 2-month follow-up |
| 2.2. Views or acceptability of design and content of programme | Progression to be agreed in conjunction with TSC based on data captured around design, content, and technical/accessibility aspects of MoodHwb. | Qualitative data from semi-structured interviews with young people and parents/carers after 2 months |

| | | |
|---|---|---|
| | Green: If changes are needed, they are minor or have been overcome over the course of the study<br>Amber: Changes needed are significant, but feasible with further development work<br>Red: Changes needed are significant and not feasible to address | Qualitative data from focus group with professionals<br><br>Questionnaire items on acceptability completed by young people and parents/carers at 2-month follow-up<br><br>Web/app usage data from Google Analytics (e.g. on technical/accessibility aspects), based on young person and parent/carer use of MoodHwb - up to 6 months after baseline |
| **3. Outcome measures for a full trial** | | |
| 3.1 Identification of primary outcomes for full trial | Discussion on the determination of primary outcome measures with TSC, including reflection on efficacy signals from candidate outcomes | Questionnaire data at 2 month follow-up<br><br>Semi-structured interviews with young people and parents/carers at 2-month follow-up<br><br>Focus group with professionals |
